# Detection of early-onset severe preeclampsia by cell-free DNA fragmentome

**DOI:** 10.1101/2024.03.22.24304708

**Authors:** Haiqiang Zhang, Longwei Qiao, Xintao Hu, Chunhua Zhang, Yu Lin, Jingyu Zhao, Xiaojuan Wu, Xiaoyan Song, Hui Tang, Ying Xue, Yang Sun, Rijing Ou, Xinxin Wang, Yan Zhang, Xin Jin, Ting Wang

## Abstract

Early-onset severe preeclampsia (EO-PE) is a distinct and highly consequential form of preeclampsia (PE), presenting significant challenges for early detection. Here, we investigated the fragmentation pattern of plasma cell-free DNA (cfDNA) in EO-PE patients. We uncovered that the nucleotide composition at the 5’ end (i.e. ends motif) of plasma cfDNA showed a unique pathological preference in EO-PE pregnancies and gestational-psychology preference in healthy pregnancies. By integrating 91 EO-PE specific motifs into a machine-learning model, we achieved accurate prediction of EO-PE development in early pregnancies. Remarkably, our model demonstrated robust performance in an independent cohort of 74 early pregnancies and 1,241 non-invasive prenatal testing (NIPT) samples with ultra-low sequencing depth. Additionally, we revealed that these PE-specific motif signatures lacked tissue specificity, originating extracellularly, and were associated with the abnormal concentration of DNA fragmentation factor subunit beta (DFFB) in EO-PE patients’ plasma. These findings establish the plasma DNA fragmentome as a non-invasive and cost-effective biomarker that can be simultaneously captured during NIPT for early EO-PE detection and provide valuable insights into cfDNA production mechanisms in preeclampsia patients.

## Introduction

Preeclampsia (PE) is one of the most serious pregnancyLJspecific complications and the leading cause of maternal and perinatal morbidity and mortality worldwide(*1*, *2*). This multisystem maternal syndrome is characterized by new-onset hypertension and maternal organ damage that occurs after 20 weeks of gestation(*3*). Among the various forms of PE, early-onset preeclampsia, occurring before 34 weeks of gestation, represents the most severe manifestation, associated with a higher incidence of maternal complications, perinatal mortality, and morbidity compared to late-onset PE(*4*, *5*). However, the complex etiology, diverse clinical manifestations, and pathophysiology of PE present significant challenges in terms of prediction, management, and prevention(*6–8*). Despite extensive efforts, clinically satisfactory biomarkers and prediction models for early detection of PE are lacking, primarily due to limited clinical availability and low predictive efficacy(*9*, *10*). Nevertheless, recent advancements in the field of circulating cell-free DNA (cfDNA) provide new opportunities for developing effective PE biomarkers. Promising results have shown that cfDNA concentration is elevated in PE and correlates with disease severity(*11*, *12*). Moreover, disparities in cfDNA coverage across promoter regions have been observed between PE and healthy pregnancies(*13*). However, further in-depth investigations and verifications are required to assess the applicability of cfDNA in early PE detection.

Recently, the study of cfDNA fragmentation patterns, known as “fragmentomics,” is an emerging field with the potential to advance our understanding of cfDNA’s biology and develop new biomarkers for clinical applications(*14*). In addition to fragment size(*15*), recent advances in this new “omics” area have identified several crucial characteristics of cfDNA fragments, including nucleosome footprints(*16*, *17*), fragment end motifs(*18*, *19*), preferred ends(*20*, *21*), jagged ends(*22*), cleavage profile(*23*) and coverage/end imbalance in regulatory elements(*24*, *25*). These characteristics are believed to preserve valuable information about the tissue of origin of cfDNA(*26*) For instance, the preferred ends of short and long plasma cfDNA are related to fetal and maternal origin(*27*). The presence of distinct characteristic end motifs in plasma cfDNA derived from cancers, placenta, and hematopoietic cells enables the monitoring of specific pathophysiological condition(*28*) Therefore, cfDNA fragmentomics shows great promise in clinical applications, particularly for noninvasive early disease detection(*29*, *30*).

In this study, we present a comprehensive analysis and comparison of end motif patterns in plasma DNA from pregnancies with early-onset severe preeclampsia and healthy pregnancies. Our investigation focuses on identifying specific sets of motifs associated with PE pathology and gestational physiology. To validate their potential for early PE detection, we assess the performance of these PE-specific motifs, and notably, our findings demonstrate robust performance even with low-coverage non-invasive prenatal testing (NIPT) data. Additionally, we explore the tissue origin of the PE-specific motif signatures, examining their relationship with cfDNA size, nucleosomal structure, and the concentrations of nucleases involved in cfDNA digestion in both EO-PE and non-EO-PE pregnancies.

## Results

### Characteristics of pregnancies in the discovery cohort

For the discovery cohort (SZ-1 cohort), we collected a total of 151 plasma samples from 140 pregnant women with early-onset PE (N=79) and without PE (N=61) at different gestational ages. Specifically, one set of plasma samples was obtained before labor during the third trimester: 16 samples were collected at the time of diagnosis of early-onset severe PE (mean gestational age, 29.9 weeks) (PT3), and 8 samples were collected before labor from healthy control women (mean gestational age, 39.7 weeks) (NT3). Another set of plasma samples was obtained during the second trimester: 71 samples were collected before the diagnosis of early-onset severe PE (mean gestational age, 17.6 weeks) (PT2), and 56 samples were collected from gestational age-matched healthy control women (mean gestational age, 19.0 weeks) (NT2). Among these subjects, 8 PE and 3 healthy pregnancies contributed plasma samples at both the second and third trimesters. (Figure 1A).

**Figure 1.**
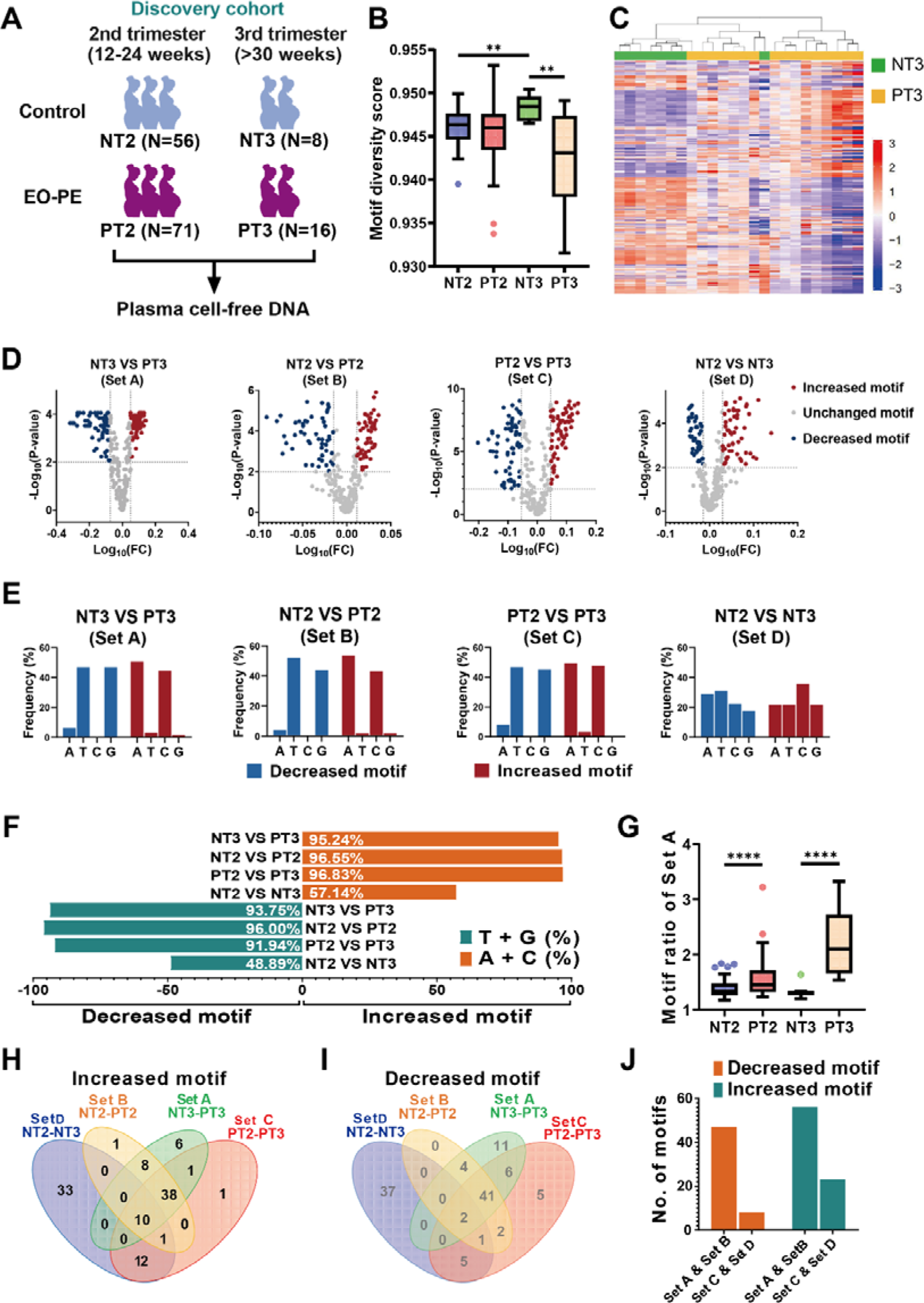
Identification of PE-specific motif pattern. (A) Sample collection in the discovery cohort. (B) Boxplot analysis of motif diversity score in plasma DNA from NT2, PT2, NT3 and PT3 groups. (C) Heatmap analysis of frequencies of 256 4mer motifs between NT3 and PT3 groups. (D) Volcano plot analysis of motifs with differential frequencies between different comparisons. The red and blue dots indicate the motifs with a P-value below 0.01 and ranked in the top 25% and bottom 25% of the fold change, respectively, which were deemed as specifically increased and decreased motifs in the latter group of the comparisons (e.g. PT3 in the comparison of “NT3 VS PT3”). Gray dots represent non-differential motifs that did not reach the thresholds. (E) Frequency of the first nucleotide at the 5’ start of all differential motifs in each comparison set. (F) The total percentage of T- and G-end motifs in the decreased motifs and the total percentage of A- and C-end motifs in the increased motif group for each group comparison. (G) Comparison of PT3-specific (identified from NT3 VS PT3, Set A) motif ratios between NT2 and PT2 groups, and between NT3 and PT3 groups. Venn diagram analysis of increased motifs H) and decreased motifs (I) among different group comparisons. (J) Number of motifs that overlapped between different group comparisons.

The demographic and clinical characteristics of all subjects are shown in Table 1. The maternal age, BMI, times of pregnancies and delivery were comparable between PE and control subjects in both 2^nd^ and 3^rd^ trimesters. Compared with control subjects (mean: 39.7 weeks), PE patients exhibited significantly lower gestational age in groups of 3^rd^ trimester (mean: 29.9 weeks) (*P*-value<0.0001, Mann-Whitney U test). The mean gestational age of PE onset in subjects collected at the 2^nd^ and 3^rd^ trimesters were 30.0 and 29.7, respectively. The maternal plasma cfDNA of group NT2, PT2, NT3, and PT3 was sequenced to a mean of 5.46 (range: 3.06 - 8.85), 5.47 (range: 2.74 - 16.3), 7.94 (range: 5.46 - 10.19) and 10.31 (range: 7.61 - 13.42) haploid human genome coverage, respectively.

**Table 1.** Demographic and clinical characteristics of pregnant women with and without EO-PE of the discovery cohort.

| Variables | Second trimester |  |  | Third trimester |  |  |
| --- | --- | --- | --- | --- | --- | --- |
|  | Controls<br>n=56 | PE<br>n=71 | P value | Controls<br>n=8 | PE<br>n=16 | P value |
| Maternal age, years | 34(25-43) | 34(24-43) | 0.779 | 34(26-38) | 33.5(27-41) | 0.916 |
| BMI, kg/m<sup>2</sup> | 23.6±3.1 | 23.9±3.5 | 0.596 | 26.0±2.6 | 28.4±4.7 | 0.230 |
| Times of pregnancies | 2.8±1.6 | 2.8±1.5 | 0.997 | 2.4±1.8 | 3.1±1.7 | 0.267 |
| Times of delivery | 0.3±0.4 | 0.3±0.5 | 0.843 | 0.4±0.5 | 0.9±0.6 | 0.086 |
| GA at sampling, weeks | 19.0±4.9 | 17.6±1.9 | 0.861 | 39.7±1.1 | 29.9±3.4 | <0.0001 |
| GA at PE was diagnosed, weeks | - | 30.0±3.0 | - | - | 29.7±3.3 | - |
| GA at delivery, weeks | - | - | - | 39.7±1.2 | 31.0±3.35 | <0.0001 |
| Natural delivery, %(n) | - | - | - | 62.5(5) | 6.25(1) | 0.003 |
| Stillbirth, %(n) | - | - | - | 0 | 6.25(1) | 0.470 |
| Male fetus, %(n) | - | - | - | 50 ( 4 ) | 28.6(4) | 0.315 |
| Birthweight, g | - | - | - | 3187.5±322.7 | 1342.7±435.0 | <0.0001 |

PE, pre-eclampsia; BMI, body mass index; GA, gestational age.
Values were given as mean±SD and median(range).
P value: The Chi-square test was used to test variables with a 0-1 distribution, such as
Natural delivery, Stillbirth, and Male fetus. The remaining continuous or discrete
variables, such as Maternal age, Times of pregnancies, and Birthweight, were
examined applying the Mann Whitney U test, with the exception of BMI, which was
evaluated with the Unpaired T test.

### Ends motifs of plasma DNA specific to PE pathology and gestational physiology

The distribution of nucleotide string at the end of plasma DNA, referred to as ends motif, was reported to embody the cfDNA fragmentation footprint(*18*). Therefore, we wonder whether the ends motif could be used to decipher the disease signatures associated with preeclampsia in circulating cell-free DNA. Accordingly, we examined the diversity of 4mer ends motifs in the plasma DNA of both control and PE groups using the motif diversity score (MDS)(*18*). The results showed that the MDS values between the control (median: 0.946, range: 0.940-0.950) and preeclampsia subjects (median: 0.946, range: 0.934-0.953) were comparable during the second trimester (*P*-value=0.107; Mann-Whitney U-test). However, during the third trimester, the MDS was significantly reduced in preeclampsia samples (median: 0.943, range: 0.932-0.949) when compared to controls (median: 0.948, range: 0.947-0.950) (*P*-value=0.003; Mann-Whitney U-test; Figure 1B), implying that preeclampsia-associated ends motif signals might be enhanced as the onset of preeclampsia approaches.

Subsequently, through the unsupervised clustering, we examined the distribution patterns of all 256 types of 4-mer motifs between the healthy and PE pregnancies in the third trimester. As shown in Figure 1C, EO-PE and healthy samples could be readily separated into two distinct clusters, suggesting the profiles of cfDNA ends motifs were highly different between pregnancies with and without EO-PE. Moreover, we compared the motif frequencies between healthy and preeclampsia pregnancies at different gestational ages, as well as between pregnancies during the second and third trimesters in preeclampsia and healthy pregnancies. We identified four sets of motifs (Log_10_(FC) ranked top and bottom 25% and Wilcox *P*-value<0.05): Set A includes 63 increased and 64 decreased motifs that exhibited a significant increase and decrease, respectively, in PT3 relative to NT3; Set B includes 58 increased and 50 decreased motifs that exhibited a significant increase and decrease, respectively, in PT2 relative to NT2; Set C includes 63 increased and 62 decreased motifs that exhibited a significant increase and decrease, respectively, in PT3 relative to PT2; and Set D includes 56 increased and 45 decreased motifs that exhibited a significant increase and decrease, respectively, in NT3 relative to NT2 (Figure 1D and Supplemental Table 1). The top 10 increased and decreased motifs of each set are shown in Supplementary Figure 1.

Notably, we observed that in both the second and third trimesters, the motifs that were significantly elevated in preeclampsia samples predominantly began with A and C (PT3: 95.24%, PT2: 96.55%), while significantly reduced motifs in preeclampsia samples mainly started with T and G (PT3: 93.75%, PT2: 96%). Such pattern was further pronounced in PT3 pregnancies compared to PT2 pregnancies (A% and C% among increased motifs: 96.83%, T% and G% among decreased motifs: 91.94%), thus demonstrating that the PE-specific motif signals become stronger as the onset of the condition approaches (Figure 1E, F). In contrast, this PE-specific signal was absent in healthy pregnancies between different gestational ages (Figure 1E). Compared with NT2, we found that in the NT3 group, the motifs starting with A and C only accounted for 57.14% of the significantly increased motifs, and the motifs began with G and T only accounted for 48.89% of the significantly decreased motifs (Figure 1F).

To quantify the signatures of preeclampsia in plasma DNA, motif ratios between the frequencies of increased and decreased motifs of Set A were calculated in PE and control pregnancies in both trimesters (See Methods). As shown in Figure 1G, the motif ratios significantly increased from PT2 (median: 1.45) to PT3 (median: 2.1) (*P*-value<0.0001, Mann-Whitney U test). Both PT3 and PT2 samples exhibited significantly increased motif ratios relative to the healthy pregnancies in the same trimester (median motif ratio of NT3: 1.31, *P*-value<0.0001; median motif ratio of NT2: 1.34, *P*-value<0.0001, Mann-Whitney U test). These results indicated that the PE-specific motifs were capable of reflecting the PE disease signals.

We further compared the consistency of differential motifs among different comparison sets. According to Fig 1H-J, 48 increased and 43 decreased motifs were co-present across Sets A, B and C, accounting for 71.65%, 84.26% and 72.8% of the motifs in Sets A, B and C, respectively. In contrast, only 10 increased and 2 decreased motifs co-existed in all sets, accounting for 9.45%, 11.11%, 9.6% and 11.88% of motifs in Set A, B, C and D. This result suggests that the motifs related to preeclampsia pathology were not shared in normal pregnancies. Moreover, 23 increased and 8 decreased motifs overlapped between Set C and Set D, which were deemed motifs altered with gestational ages, accounting for 24.8% and 30.69% of motifs in Set C, and D, respectively (Figure 1H-J). These shared motifs were mostly gestational physiology-related signals that coexisted in both preeclampsia and healthy samples.

### Correlation between gestational age and PE-specific motif abundance

In the Discovery cohort, 3 pregnant women in the control group and 8 pregnant women in the PE group contributed samples during both 2nd trimester and 3rd trimesters. These samples allowed us to further investigate how the PE-specific motif signals (i.e. frequencies of motifs in set A) changed as the pregnancy progressed. As shown in Figure 2A, most of the PE samples displayed enhanced trends for increased and decreased Set A PE-specific motifs in PT3 compared with PT2. While the PE-specific motif frequencies remained consistent between NT3 and NT2 samples (Figure 2B), indicating the absence of PE signals in healthy pregnancies. In order to quantify the enhancement of motif patterns, we calculated the accumulated deviations between motif frequencies in samples collected from 2^nd^ and 3^rd^ trimesters for each pregnant woman. We termed this value as “motif distance”. According to Fig 2C, the PE samples showed dramatically higher motif distances than the control samples. 75% (6/8) of the PE pregnancies showed enhanced motif patterns (i.e. motif distance >0), but none of the healthy pregnancies exhibited this enhancement.

**Figure 2.**
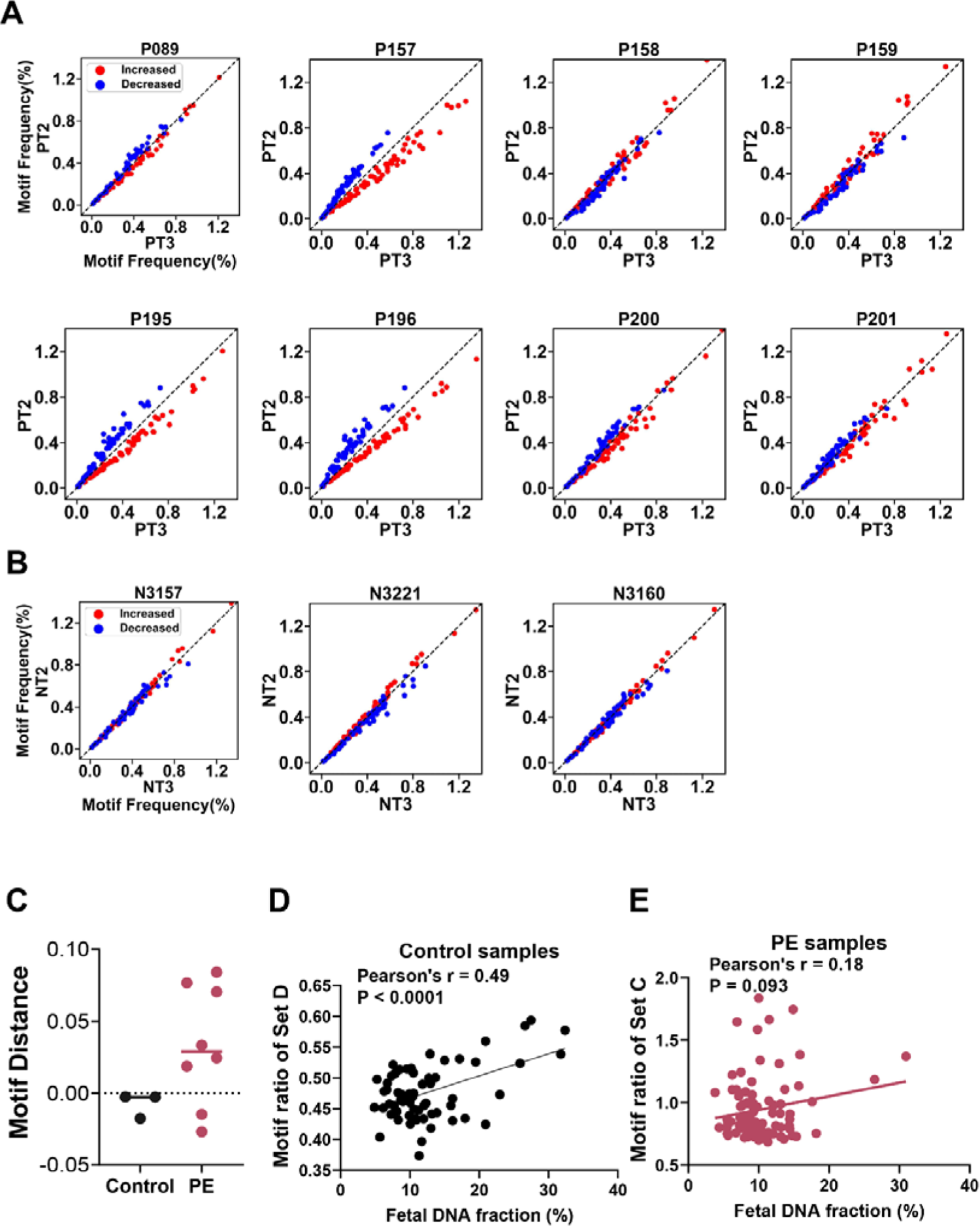
Motif distributions in pregnant women with different gestational ages. (A) The frequencies of PE-specific motifs in plasma DNA samples collected from the same PE pregnant woman during her second and third trimesters. (B) The frequencies of PE-specific motifs in plasma DNA samples collected from the same healthy pregnant woman during her second and third trimesters. (C) Boxplot analysis of motif distance between plasma DNA of second and third trimesters in control and PE samples. (D) Correlation between NT3-specific (Set D) motif ratio and fetal DNA fraction in maternal plasma of healthy pregnant women. (E) Correlation between PT3-specific (Set C) motif ratio and fetal DNA fraction in maternal plasma of pregnant women with PE.

### Correlation between fetal DNA fraction and gestational physiology-related motif abundance

To assess the gestation-physiology signals in PE and healthy pregnancies, we analyzed the correlation between set D motif ratios and fetal DNA fraction in maternal plasma in samples of the two groups. The motif ratios were observed to be significantly correlated with the fetal DNA fractions in healthy samples (Pearson r= 0.49, *P*-value<0.0001), further consolidating that the motifs in set D were mostly gestation related and carry strong physiology signals (Figure 2D). However, the correlation between Set C motifs ratio and the fetal DNA fraction in PE samples was not significant (Figure 2E). This weak correlation in PE samples might be due to the confounder of abundant PE pathology signals (i.e. PE-specific motifs in set C) carried.

### Early identification of EO-PE patients by PE-specific motif ratio

As the cfDNA ends motifs of PE patients showed distinct PE signatures, we wondered whether these PE-specific motifs could be used as diagnostic biomarkers in the early detection of preeclampsia during pregnancy. Hence, we further recruited an independent pregnancy cohort containing early-onset severe PE (N=26, gestational age: 17.5±2.9 weeks) and gestation-age-matched healthy (N=48, gestational age: 17.8±2.8 weeks) pregnancies in the 2^nd^ trimester as a validation cohort (Supplemental Table 2) in the same hospital (SZ-2 cohort). The plasma samples were processed with the same protocol and platform as the discovery cohort, with mean sequencing depths of 19.03 (range: 9.73 - 87.84) and 22.61 (range: 8.32 - 103.27) for control and EO-PE subjects, respectively. Motif ratios based on PE-specific motifs of Sets A and B from the discovery cohort were analyzed in this validation cohort (See Methods). As shown in Figure 3A and B, the median of motif ratios based on set A and B in EO-PE samples were both significantly higher than healthy samples (median set A motif ratio: 1.45 vs 1.30, *P*-value<0.0001; median set B motif ratio: 1.28 vs 1.16, *P*-value<0.0001, Mann-Whitney U test), in line with the observations in the discovery cohort (Figure 1G), which validated the robustness the PE-specific motifs. ROC curve analysis was further employed to assess the classification performance between PE and healthy pregnancies in this validation cohort by motif ratios. The AUC was 0.78 [95% CI:0.65-0.92] for the PE determination based on motifs of set A, which was comparable to the use of motifs of set B (AUC:0.77 [95% CI: 0.63-0.90]) (Figure 3C).

**Figure 3.**
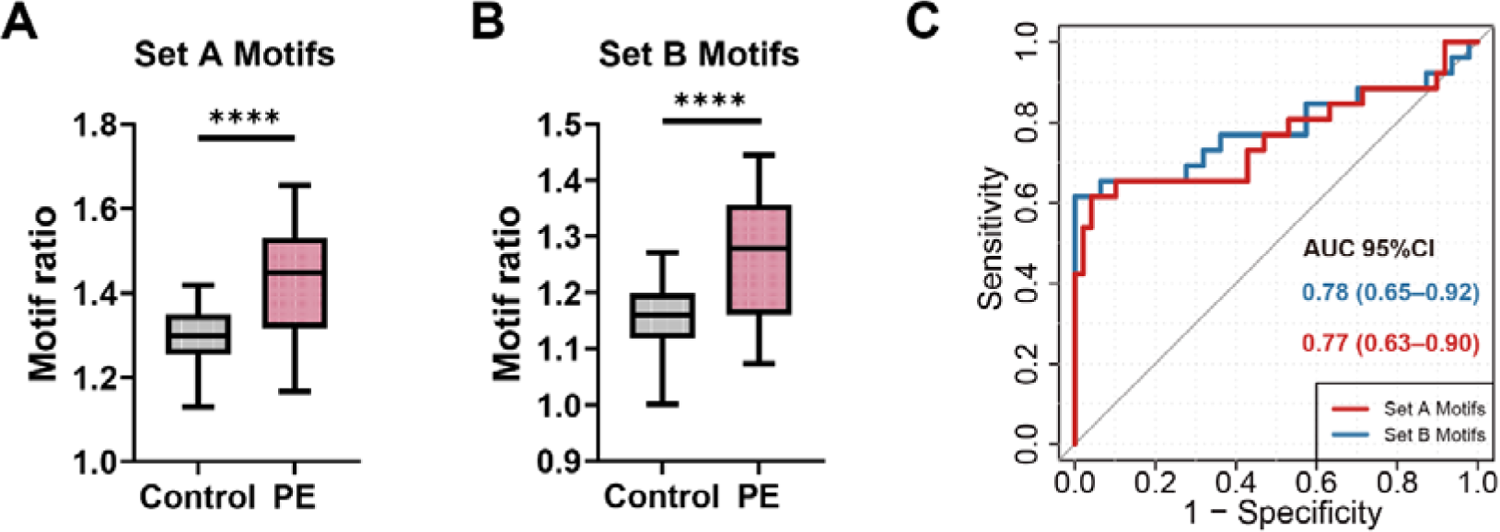
PE-specific motif analysis in plasma DNA of SZ-2 validation cohort. Boxplot of the motifs ratio in PE and control samples based on the PE-specific motifs of Set A (A) and Set B (B). (C) ROC curve analysis between PE and control groups with the use of Set A and Set B motifs ratios.

### Early identification of EO-PE patients based on machine learning model

Since a single PE-motif ratio demonstrated the potential for PE identification, we aimed to develop a machine-learning model based on PE-specific motifs to maximize the PE prediction performance. As shown in Figure 4A, first, we extracted 91 PE-specific motifs shared among Sets A, B and C of the discovery cohort as the model features. As the motifs were mined and validated in the pair-end sequencing data with high sequencing depth, to assess the applicability and robustness of these motif biomarkers in samples with different sequencing platforms, protocols, and ultra-low sequencing depth, we further collected 2,481 sequencing samples from pregnancies (Healthy pregnancy (HP): n=2445, EO-PE pregnancy: n=36) receiving non-invasive pregnant testing (NIPT). From this dataset, we randomly selected 1,222 healthy samples and 18 EO-PE samples (NIPT-1 dataset) and combined them with the discovery cohort as training samples. The remaining NIPT samples (NIPT-2 dataset) were used as testing samples. A support vector machine (SVM) model for PE detection was trained and determined through the 10-fold cross-validation (See Methods). The prediction model was applied to the SZ-2 cohort and NIPT-2 dataset for the EO-PE detection.

**Figure 4.**
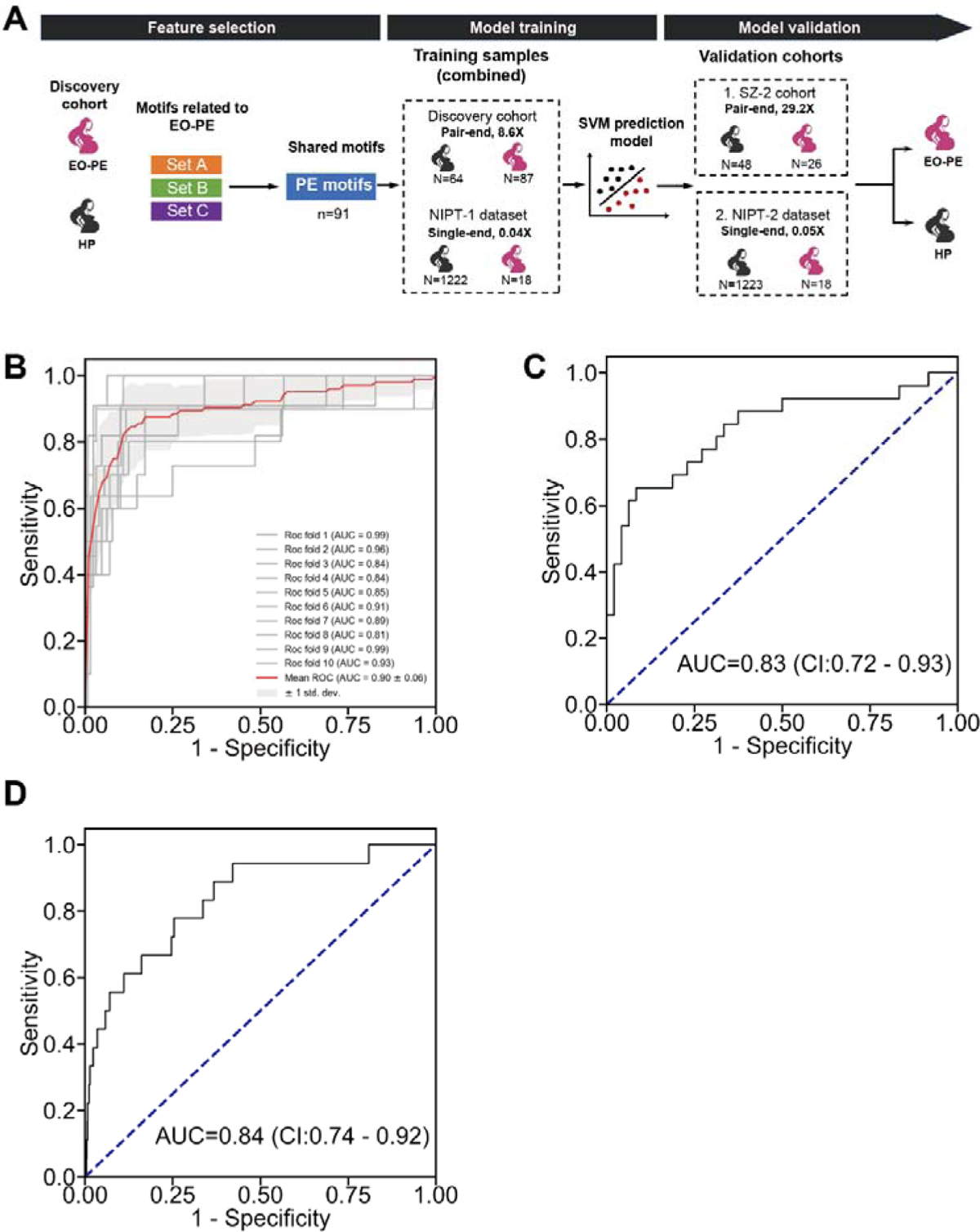
PE prediction in SZ-2 and NIPT cohort based on SVM model (A) Illustration of PE prediction using PE-specific motifs by SVM model. (B) ROC curve analysis between PE and control groups in pooled training samples based on the prediction model using 91 PE-specific motifs and the 10-fold cross-validation ROC curve of EO-PE detection in SZ-2 cohort (C) and NIPT-2 sample set (D) based on PE prediction model.

As a result, we obtained AUC values of 0.90 ± 0.06 for determining EO-PE patients in the training sample set (Figure 4B). Moreover, we achieved a high AUC of 0.83 [95% CI: 0.72-0.93] for EO-PE detection in the SZ-2 validation cohort (Figure 4C). Notably, even with ultra-low sequencing depth (0.05X) and single-end sequencing mode, we obtained a high AUC of 0.84 [95% CI: 0.74-0.92] for the NIPT-2 sample set (Figure 4D).

### Tissue-of-origin of cfDNA carrying PE-specific motifs in preeclampsia pregnant woman

The cell-free DNA in maternal plasma could be released by the fetus and the mother. To explore whether the PE motif signals were specifically origin from the fetus (i.e. placenta) or the mother (i.e. predominantly the hematopoietic cells), we reanalyzed the plasma DNA of one healthy and one PE pregnant woman with matched sequencing data of maternal and paternal buffy coat from our previously published paper(*31*) (Figure 5A). Fetal- and maternal-derived cfDNA were determined by the parental specific alleles (See Methods). We categorized the plasma DNA into four origin groups: (1) PE-mother and (2) PE-fetus that differentiated from the PE individual, (3) Control-mother and (4) Control-fetus that differentiated from the healthy individual (Figure 5A).

**Figure 5.**
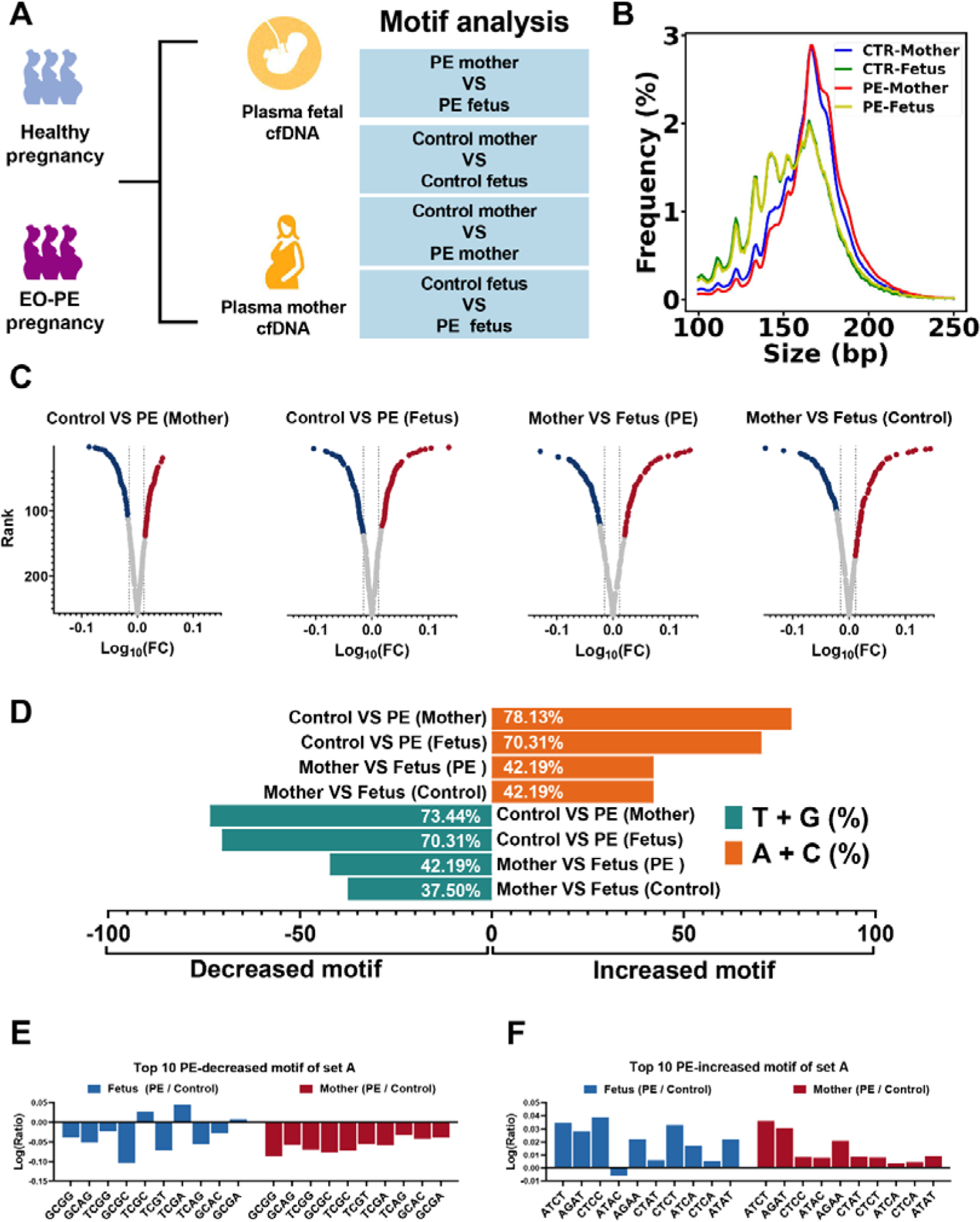
Ends motif of fetal- and maternal-derived cfDNA in a healthy and a PE pregnant woman. (A) Illustration of motif analysis in fetal and maternal cfDNA in pregnant women with and without PE. Four sets of motif comparisons were performed between fetal- and maternal-derived cfDNA and between PE and healthy pregnant women. (B) Size profiles of fetal- and maternal-derived cfDNA in healthy and PE pregnant women. (C) Volcano plot analysis of motifs with differential frequencies between different comparisons. Y-axis indicates the rank of the motifs according to their frequency in descending order. Red and blue dots indicate the motifs ranked in the top 25% and bottom 25% of the fold change, respectively, which were deemed as specifically increased and decreased motifs in the latter group of the comparisons. Gray dots indicate those motifs below the thresholds, which were deemed as non-differential motifs. (D) The total percentage of T- and G-end motifs in the decreased motifs and the total percentage of A- and C-end motifs in the increased motif group for each group comparison. Logarithmic scale representation of the ratio between frequencies of top 10 PE-decreased motifs (E) and top 10 PE-increased motifs (F) of set A (based on the discovery cohort) in plasma fetal- and maternal-derived cfDNA of PE and healthy pregnant women.

As shown in Figure 5B, the plasma DNA originating from the fetuses was significantly shorter than mother-originated ones. Besides, the PE-mother derived cfDNA was observed to be longer than the cfDNA from the control-mother. To investigate whether the identified PE motif pattern is specific to either the mother or the fetus, we conducted a comparative analysis of motif frequencies between different sources of cfDNA, including (1) fetus-derived cfDNA from PE and control pregnancies, (2) maternal-derived cfDNA from PE and control pregnancies, as well as mother-derived and fetus-derived cfDNA within the (3) PE patient and (4) healthy pregnancy (Figure 5A and C). We calculated the log_10_ (Fold change) of 256 motifs in terms of frequency within each of the four group comparisons and ranked the motifs based on the |log_10_ (Fold change)|. We took the top and bottom 25% of motifs with positive log_10_ (Fold change) and top 25% of motifs with negative log_10_ (Fold change) values as the differentially increased and decreased motifs in the latter pregnant woman of the comparison (e.g. PE in the comparison of Control VS PE) (Figure 5C). The motif pattern specific to preeclampsia, characterized by an enrichment of A- and C-started motifs and a reduction of G and T motifs, was consistently observed in both fetal-derived and maternal-derived cfDNA of PE pregnancy compared to those of the healthy pregnant woman (Supplemental Figure 2 and Figure 5D). Notably, the A- and C-started motifs accounted for 78.13% and 70.31% of the differentially increased motifs, while the T- and G-started motifs accounted for 73.44% and 70.31% of the differentially decreased motifs in PE-mother and PE-fetus derived cfDNA, respectively, when compared with those in healthy pregnancy. However, this motif pattern was absent in the comparison between fetus-derived cfDNA and mother-derived cfDNA in either PE or control subjects. Among the increased motifs, the A and C-started motifs only account for 42.19% and 42.19% in the fetus-derived cfDNA compared to the mother derived in PE and control subjects, respectively. Among the decreased motifs, the G and T-started motifs only account for 42.19% and 37.50% in the fetus-derived cfDNA compared to the mother derived in PE and control subjects, respectively (Figure 5D).

Moreover, we compared the motif ratio of the top 10 increased and top 10 decreased motifs of set A between fetal cfDNA from both PE and healthy pregnancies, and maternal cfDNA from both PE and healthy pregnancies. As shown in Figure 5E, for PE-decreased motifs, 7 out of 10 in PE-fetus derived cfDNA and all of them in PE-mother derived cfDNA showed consistently decreased patterns compared with that origin in the healthy individual. Meanwhile, for PE-increased motifs, 9 out of 10 in PE-fetus derived cfDNA and all of them in PE-mother derived cfDNA showed consistently increased patterns compared with that origin in healthy individuals (Figure 5F). These results demonstrated that the PE-specific motif pattern co-existed in both the fetal and maternal released DNA in plasma. Therefore, as the motif differences observed in both the mother and fetus with preeclampsia exhibit congruent patterns, we speculated that the fragmentation preference producing the PE-specific motifs was non-tissue-specific and primarily attributable to extracellular DNA molecule degradation, which might be influenced by nucleases in the plasma, and shared the same pattern in both the mother and fetus.

### Size and nucleosome footprint of cfDNA carrying different ending nucleotide

Ends motifs were generated from the fragmentation of cfDNA in plasma, which was influenced by both the nucleosome binding patterns(*27*) and the activity of nuclease(*32*). Thus, to explore the potential fragmentation factors leading to the preference of motifs in PE patients, we analyzed the size of DNA molecules and the occurrence of cfDNA ending sites in relation to nucleosome centres. For each DNA fragment, the upstream and downstream 5’ ends were labelled as the U-end and D-end, respectively (Figure 6A). We randomly selected 8 samples from each of the groups in the discovery cohort and pooled them to represent that group.

**Figure 6.**
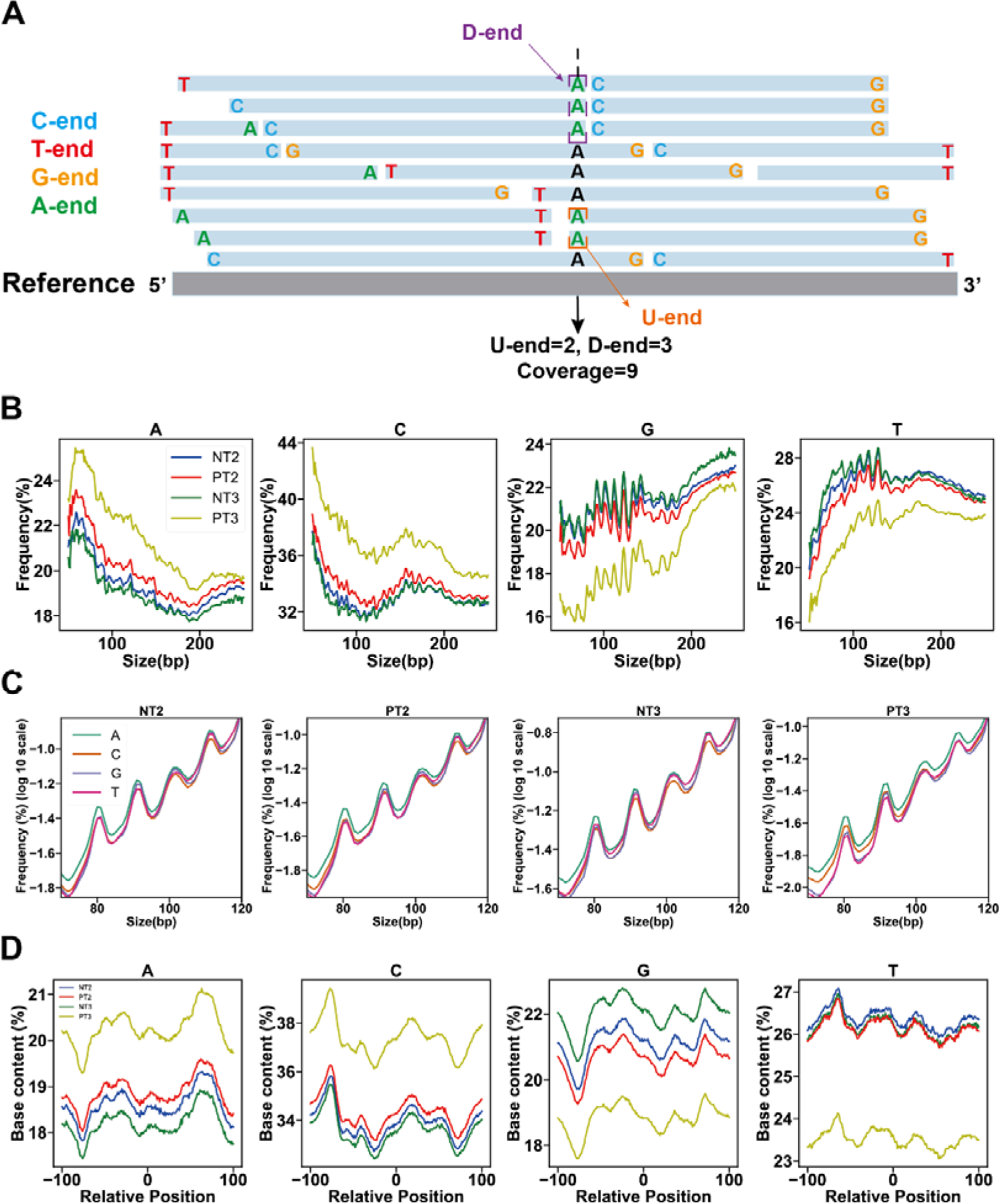
Size and nucleosome footprint analysis for cfDNA carrying different U-ends. (A) Illustration of ends occurrence and coverage of cfDNA with different ends across the genome. (B) Composition of plasma cfDNA with different U-ends in NT2, PT2, NT3 and PT3 groups at each of the cfDNA lengths between 50 and 250bp. The frequency of cfDNA at each individual size was determined by calculating the number of cfDNA molecules with a specific ending nucleotide and normalizing it to the total number of cfDNA molecules with the same size. (i.e. in each size, the summed frequency of cfDNA with A, T, C and G ends was 100%). (C) Size profiles of cfDNA with different U-ending nucleotides across 50 and 120bp in different groups (i.e the frequency of cfDNA with particular ending nucleotides at each individual cfDNA length was normalized by the amount of cfDNA with the same ending nucleotides between 50 and 120bp). (D) Composition of cfDNA with different U-ending nucleotides at each genomic location relative to nucleosome structure in NT2, PT2, NT3 and PT3 groups. The occurrence of each type of cfDNA ends in each relative position of nucleosome was normalized by the total number of ends in this location.

First, we analyzed the content of cfDNA molecules carrying different nucleotides (i.e. A, T, C and G) in the U-end (Figure 6B) and D-end (Supplemental Figure 3A) within the size range of 50-250bp in all groups. As shown in Figure 6B, the PT2 and PT3 groups showed consistently higher contents of cfDNA with A- and C-end and consistently lower contents of cfDNA with G- and T-end compared with control groups. Additionally, cfDNA with different ending nucleotides displayed dramatically different proportion profiles. cfDNA with C-end showed the highest proportion (i.e. >30%) among all groups across all sizes and displayed a predominant peak around 166bp, which was reminiscent of the length of DNA wrapping the nucleosome core, indicating that the nuclease cutting on C-end DNA may primarily occur on the link DNA of nucleosome and contributed the majority of cfDNA. For short cfDNA below 100bp, the proportions of cfDNA with A-end and C-end were initially higher and decreased gradually. Conversely, for cfDNA ending with G and T, the proportions of such cfDNA progressively increased below 150bp and exhibited a strong 10-bp periodicity. These findings suggest that the components of cfDNA with different ends at each size exhibit distinct preferences and fluctuate accordingly.

Figure 6C displays the size profiles of cfDNA with different U-end nucleotides between 50bp and 120bp. The cfDNA with A-end exhibited a higher proportion of short DNA (e.g. <120bp) among all groups. The cfDNA with C-end further showed a slightly higher proportion in the PT2 group and a distinctly higher proportion in the PT3 group. Similar patterns were also observed in cfDNA with different D-end nucleotides (Supplemental Figure 3B). These results indicated that the PE-specific cleavage signals existed in cfDNA across various sizes.

To explore whether the cfDNA with different ends would have different cutting preferences in relation to nucleosome structure, we mapped the ending sites of cfDNA with A, T, C, and G ends to the upstream and downstream regions of the nucleosome core. Figure 6D illustrates the content of cfDNA with different U-ends in each location relative to the nucleosome core. The cfDNA with different ends showed distinct proportions across nucleosome structures. For example, the cfDNA with A and G ends showed troughs around 75bp upstream to the nucleosome core, while the cfDNA with C and T ends showed peaks at this location. The PE and control groups showed consistent waveforms in the distributions of the cfDNA with the same ending nucleotide. However, compared with control groups, the component of A-end and C-end cfDNA were elevated in PT3 and PT2 groups, the component of G-end and T-end cfDNA were reduced in PT3 and PT2 groups, across the 200bp regions around the nucleosome core. The cfDNA distribution patterns in D-end are shown in Supplemental Figure 3C. These results show that cfDNA with different ending nucleotides undergoes distinct cutting patterns across nucleosome structures, whereas PE cleavage patterns can occur throughout the entire nucleosome structure rather than being limited to specific regions.

### Nuclease levels in plasma DNA of healthy and PE pregnancies

The above findings revealed that the PE cleavage signals were present in all cfDNA sizes and in regions surrounding the nucleosome core. We speculated that the activity of nucleases in plasma might be altered and thus lead to systematically different dynamics of fragmentation patterns. Figure 7A shows the cfDNA concentrations in the plasma of different groups. The patients with preeclampsia were observed to have significantly higher cfDNA levels than the healthy pregnancies at both 2^nd^ (median: 41.2 ng/μl vs 30 ng/μl, *P*-value<0.0001, Mann-Whitney U test) and 3^rd^ (median: 52.4 ng/μl vs 41.8 ng/μl, *P*-value=0.0001, Mann-Whitney U test) trimesters, indicating the nuclease activity and cfDNA generation might be more pronounced in PE patients in comparison to healthy pregnant women.

**Figure 7.**
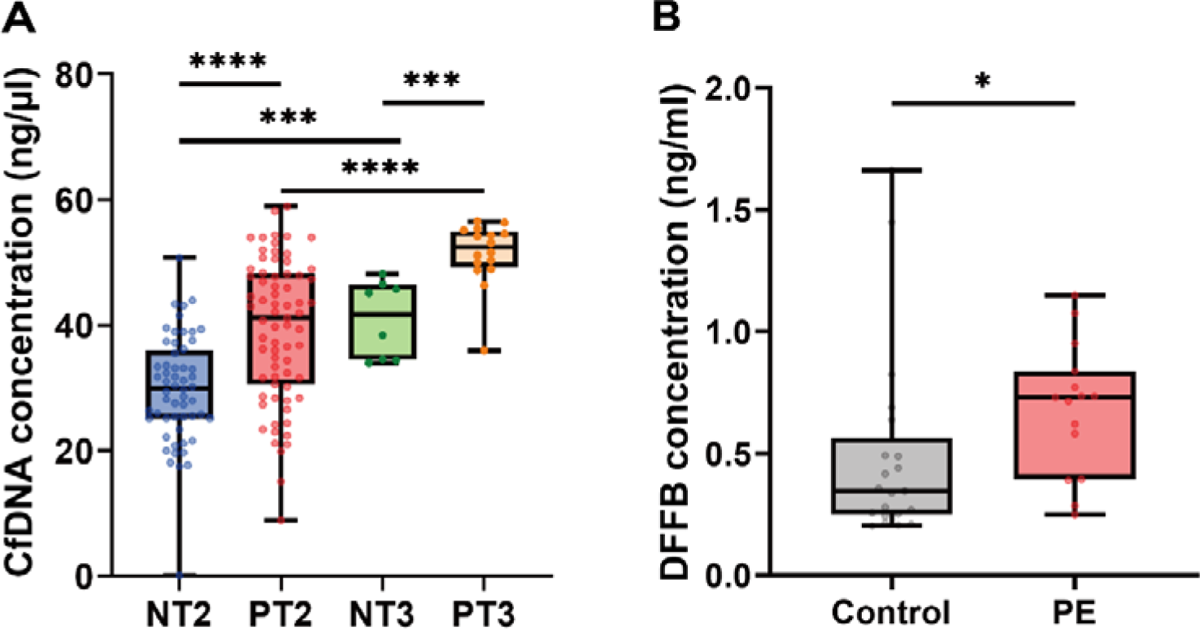
(A) cfDNA concentration in the plasma of NT2, PT2, NT3 and PT3 groups. (B) The concentrations of DFFB in the plasma of NT2, PT2, NT3 and PT3 groups.

Previous studies(*32*) have implicated the involvement of nuclease DFFB and Deoxyribonuclease I (DNASE1) in cfDNA fragmentation in plasma. Specifically, DFFB has been shown to preferentially generate cfDNA with A ends, while DNASE1 is associated with the production of cfDNA with T-ends. Thus, we further quantified the concentration of nuclease DFFB and DNASE1 in the plasma of 21 healthy and 15 EO-PE pregnancies during the 3^rd^ trimester. The median levels of DFFB were approximately two-fold higher in PE subjects (median: 0.73 ng/ml, range: 0.25 ng/ml −1.15 ng/ml) compared to healthy subjects (median: 0.36 ng/ml, range: 0.20 ng/ml −1.66 ng/ml) (*P*-value=0.011, Mann-Whitney U test) (Figure 6F). This increase in DFFB levels might contribute to the higher proportion of cfDNA with A ends observed in PE pregnancies. Besides, we also observed significantly increased levels of DNASE1 in PE patients (median: 4.00 ng/ml, range: 1.71 ng/ml −8.45 ng/ml) compared with healthy individuals (median: 1.98 ng/ml, range: 0.86 ng/ml −9.22 ng/ml) (*P*-value=0.0110, Mann-Whitney U test) (Supplemental Figure 4).

## Discussion

In this study, we investigated the end motif patterns in the plasma DNA of pregnancies with and without early-onset severe preeclampsia in different trimesters. Our findings revealed that these end motifs exhibit distinct pathological signatures in preeclampsia patients, while displaying physiological signatures in healthy pregnant women.

By leveraging the PE-specific motifs identified in the discovery cohort, we successfully validated their utility as a highly effective biomarker for the early detection of EO-PE in another independent cohort processed with a similar protocol. Remarkably, incorporating the PE-specific motifs into a predictive model enabled us to accurately identify PE patients even in samples from the NTPT group, which underwent disparate protocols, including variations in library preparation, sequencing platforms, ultra-low sequencing depth (∼0.05X), and sequencing mode (35bp single-end read). These findings reinforce the robustness and versatility of the PE-specific motifs in early PE detection and highlight their potential for integration into the current non-invasive prenatal testing protocol.

By deciphering the motifs in maternal- and fetal-derived cfDNA in maternal plasma, we revealed that the PE-specific motif preference was likely generated within the plasma rather than in the somatic tissue (e.g. placental). Furthermore, through dissecting the component of cfDNA with different ending nucleotides at varying lengths and nucleosome structures, we uncovered that the fragmentation process leading to the formation of PE-specific motifs was enhanced across various sizes and the entirety of nucleosome-wrapped cfDNA. Specifically, the fragmentation-generating ends with nucleotides A and C showed systematic enhancement. This led us to hypothesize that the DFFB nuclease(*32*), known for its preference in generating cfDNA with A ends, might be involved in generating the specific motif pattern associated with PE. Supporting this hypothesis, we have observed a significant elevation in the concentration of DFFB in PE patients, implying the potential role of DFFB nuclease in the etiopathology of preeclampsia. Additionally, we observed a significant increase of DNASE1 in the plasma of PE patients compared to healthy pregnancies. Previous studies(*32*) have suggested that DNASE1 is prone to generating cfDNA molecules ending with T. However, in our study, we observed a reduced frequency of cfDNA ending with T in the plasma of PE patients, leading to a contradiction. One possible explanation for this discrepancy is that a published research by Cheng et al(*33*) has indicated that DNASE1 may not be a major nuclease responsible for DNA digestion in plasma. Therefore, while the concentration of DNASE1 increases in plasma, its impact on DNA fragmentation may be negligible or counterbalanced by the activity of DFFB. Instead, DNASE1 has been shown to play a major role in cfDNA fragmentation in urine(*34*). Exploring the motifs present at cfDNA ends and DNASE1 activity in urine, along with its relationship with the onset of preeclampsia, would be a worthwhile avenue for further research.

Recently, using single-molecule sequencing, Yu et al(*35*). also reported a preference of end motifs in the plasma DNA (a significant amount of long cfDNA, e.g., >500bp) of PE patients in the third trimester. Their findings, however, differ from our study in terms of the specific motif preferences observed. In our study, we observed that the majority of increased motifs in PE patients were characterized by ends with A and C (e.g., >95%), while the majority of decreased motifs in PE patients were characterized by ends with T and G (e.g., >91%). In contrast, Yu et al. found that motifs with reduced frequencies in preeclampsia predominantly ended with G or A, while those with increased frequencies all ended with T or C. This inconsistency suggests that the long DNA fragments captured by single-molecule sequencing may originate from different fragmentation patterns compared to the short cfDNA fragments sequenced by second-generation sequencing. These variations in fragmentation patterns could potentially contribute to the observed differences in motif preferences.

Despite the valuable insights gained from our study, there are certain limitations that need to be addressed in future research. Firstly, our discovery cohort consisted only of early-onset preeclampsia patients. Thus, a larger cohort containing multiple types of preeclampsia collected from multiple centers is required to explore the applicability of our findings to other types of preeclampsia. Another limitation of our study is the lack of multiple sampling for the majority of pregnancies. Only 24 pregnant women were sampled twice in our study, which limits the exploration of the earliest time for preeclampsia detection based on motif biomarkers. Including multi-point sampling in future studies would also be highly valuable in monitoring the progression of PE by capturing dynamic changes in the abundance of PE-specific motifs.

In summary, our study highlights the potential of cfDNA end motif as a promising non-invasive biomarker for early detection of early-onset severe preeclampsia in pregnancy. Additionally, we have provided significant insights into the mechanisms involved in the generation and release of cfDNA from preeclampsia patients. By extending the clinical applications of cfDNA fragmentomics, our findings offer a valuable approach for the fast and cost-effective detection of pregnancy complications.

## Methods

### Statistics

We utilized Mann-Whitney U-test (Wilcoxon Rank Sum Tests) and the Kruskal-Wallis test to evaluate differences in continuous variables among different groups. For the comparison of categorical variables, we employed the Chi-square test or Fisher’s exact test. Pearson’s test was used to assess correlation. P-values below 0.05 were considered statistically significant. P-values less than 0.05, 0.01, 0.001, and 0.0001 were denoted by *, **, ***, and ****, respectively. We conducted the statistical analysis using R software (version 3.6.1).

### Study approval

Our study was approved by the Institutional Review Board of BGI (BGI-IRB 20040-T1) and the Institutional Review Board of Suzhou Municipal Hospital (K-2019-019-K01).

### Patients and samples

For the discovery cohort (SZ-1 cohort), we recruited 140 singleton pregnancies including healthy pregnancies and those diagnosed with preeclampsia, during the 2nd and 3rd trimesters at the Suzhou Municipal Hospital, Jiang Su, China. For the validation cohort (SZ-2 cohort), 26 PE and 48 healthy pregnancies, all in the 2nd trimester, were further recruited at the same hospital.

For the NIPT cohort, the sequencing data of NIPT from 36 pregnancies who developed early-onset severe PE and 2,445 healthy pregnancies accepted NIPT between January 2016 and March 2021 were obtained from the Suzhou Municipal Hospital, Jiang Su, China. The plasma cfDNA was sequenced by Illumina Hiseq sequencing platform in a 35bp single-end format, resulting in a median of 3,497,067 sequencing reads (range: 1,060,392-11,201,854). From these samples, we randomly selected 1,222 healthy samples and 18 EO-PE samples to be included in the training dataset for the PE prediction model. The remaining samples were allocated for testing the EO-PE prediction.

To quantify nuclease concentration in maternal plasma, we collected plasma samples from 21 healthy pregnancies (8 samples were sequenced in the discovery cohort) and 15 EO-PE pregnancies (12 samples were sequenced in the discovery cohort) at the same hospital.

All participants involved in the study provided written informed consent. This study was approved by the Ethics Committee of Suzhou Municipal Hospital (K20190019H01) and the Institute Review Board of BGI (BGI-IRB 20040).

### Sample collection and processing

Peripheral blood samples were collected from each subject using EDTA tubes. The blood samples were centrifuged within 4 hours of collection at 1,600 × g for 10 minutes at 4°C. Subsequently, the supernatant was isolated and centrifuged again at 16,000 × g for 10 minutes at 4°C. This process yielded 5ml of plasma.

### Sequencing Library Preparation

CfDNA was extracted from the plasma samples using the MagPure Circulating DNA Mini KF Kit (Magen, MD5432-02). DNA libraries were prepared using MGIEasy Cell free DNA library preparation reagent kit (MGI, 1000007037) according to the manufacturer’s instructions. cfDNA concentration was measured by Qubit dsDNA HS Assay Kit (Q32854, Invitrogen).

### Quantification of Nuclease levels in plasma

For the quantification of human DFFB and DNASE1 levels in plasma, we used commercial enzyme-linked immunosorbent assay (ELISA) kits purchased from Jianglai Industrial (catalog number JL10215) and CUSABIO (catalog number CSB-E09068h). The assay values ranged from 0.15 ng/mL to 10 g/mL and 1.56 ng/mL to 100 ng/mL, respectively. The principle of the kit is based on the indirect sandwich ELISA technique, which utilizes a capture antibody and a biotin-labelled detection antibody for capture and detection purposes, respectively.

### Sequencing and Alignment

The DNA libraries were sequenced using the DNBSEQ platform (MGI) in a 100bp × 2 paired-end format. Fastp (v0.20.1)(*36*) was employed to remove adaptor sequences and eliminate low-quality reads (i.e., quality score < 20) from the raw sequencing data. Subsequently, Minimap2 (v2.2.11) was used to align the clean reads to the human reference genome (GRCh38/hg38). PCR duplicates were filtered out using Biobambam2 (v2-2.0.87)(*37*). Reads with more than three mismatches or aligning to multiple locations were filtered. only paired-end reads that aligned to the same chromosome with the correct orientation and had an insert size of ≤600 bp were retained for downstream analysis.

### Motif analysis

The 4-nucleotide sequence located at the 5’ end of plasma DNA fragments is previously defined as the ends motif(*18*). Motif frequency was computed by normalizing the count of this specific motif against the total count of all 256 types of 4mer motifs. The motif diversity score based on the entropy principle, as described before(*18*), was employed to gauge the uniformity of motif distribution. A low MDS value indicated a skewed distribution, while a high MDS value indicated a more uniform distribution of ends motifs in terms of their frequencies. Additionally, the motif ratio was determined as the ratio between the accumulated frequencies of increased motifs and decreased motifs.

### Ends analysis

cfDNA molecules possess an upstream (U) and a downstream (D) 5’ endpoint, determined by their coordinates on the human reference genome. We refer to these endpoints as the U-end and D-end, respectively. The U-end nucleotide can be one of the A/T/C/G bases, leading to the classification of cfDNA into UA/UT/UC/UG fragments. Similarly, the D-end nucleotide categorizes cfDNA into DA/DT/DC/DG fragments. To achieve a high sequencing depth of cfDNA for deep investigation, we randomly selected 8 samples from each group in the discovery cohort and pooled them to represent the group. We analyzed and compared the size profiles of cfDNA belonging to different categories.

Meanwhile, the occurrence of the ending sites of cfDNA from different categories was interrogated across the 200bp region surrounding each of the nucleosome cores determined in the plasma DNA from Synder et al‘s paper (Figure 6A)(*17*). By accumulating and normalizing the cfDNA ends relative to the nucleosome cores within this 200bp region, we determined the composition of cfDNA with different end types at each location of this region, as well as the relative frequencies of cfDNA with a specific end type across the region.

### Analysis of fetal- and maternal-derived DNA

The sequencing data of plasma DNA and paired parental genomic DNA of a healthy and a preeclampsia pregnancy were obtained from our previous study(*31*). The parental informative single-nucleotide polymorphisms (SNPs) where the mother was homozygous (AA) and the fetus was heterozygous (i.e. fetus: AB, deduced from father: AB/BB) were used for the differentiation of fetal- and maternal-derived cfDNA in maternal plasma. We determined fetal-derived cfDNA based on those molecules carrying fetal-specific alleles (B). Meanwhile, as the majority of cfDNA in maternal plasma is mother released, the molecules carrying shared alleles (A) were deemed as predominately maternal-derived.

### Development of Machine-learning model for EO-PE detection

We identified 91 differential motifs that were shared among Sets A, B, and C in the discovery cohort and utilized them as model features. The training dataset consisted of all samples from the discovery cohort, along with a random selection of 1,222 control samples and 18 EO-PE samples. To train our EO-PE prediction model, we employed the Support Vector Machine (SVM) algorithm and performed 10-fold cross-validation. The hyperparameters and model that yielded the highest AUC value in PE classification within the training sample set were selected as the final EO-PE prediction model. We then evaluated the performance of the developed EO-PE prediction model by applying it to the validation sets of the SZ-2 cohort and the NIPT-2 dataset.

## Author contributions

Conception and design, HQ.Z., X.J., T.W.; sample collection and processing, LW.Q., CH.Z., XJ.W., XY.S., H.T., Y.X., RJ.O., XX.W., Y.Z.; analysis and interpretation of data, HQ.Z., LW.Q., XT.H., Y.L., JY.Z., Y.S., X.J., T.W.; writing – original draft, HQ.Z., LW.Q., XT.H., Y.L., JY.Z.; writing – review & editing: HQ.Z., LW.Q., XT.H., X.J., T.W.; All authors reviewed the manuscript and approved the final revision.

## Data Availability

All data needed to evaluate the conclusions in the paper are present in the paper and/or the Supplementary Materials. The key raw data have been deposited onto the China National GeneBank DataBase (CNGBdb) with accession number CNP0003056.

https://db.cngb.org/search/project/CNP0003056/

## Acknowledgement

We would like to thank China National GeneBank (CNGB) for the technical support.

---

**Table S1.** List of differential motifs from different comparisons in the discovery cohort.

**Table S2.** Demographic and clinical characteristics of pregnant women with and

**Figure S1.** Top 10 motifs with specific increased frequencies (red) and top 10 motifs with specific decreased frequencies (blue) in different motif sets identified in the discovery cohort.

**Figure S2.** Frequencies of motifs with different starting nucleotides among the increased motifs (red) and decreased motifs (blue) of (A) maternal-derived cfDNA of the PE pregnancy compared to those of the healthy pregnancy, (B) fetal-derived cfDNA of the PE pregnancy compared to those of the healthy pregnancy, (C) fetal-derived cfDNA compared to maternal-derived cfDNA of the PE pregnancy, (D) fetal-derived cfDNA compared to maternal-derived cfDNA of the healthy pregnancy.

**Figure S3.** Size and nucleosome footprint analysis for cfDNA carrying different D-ends. (A)Composition of plasma cfDNA with different D-ends in NT2, PT2, NT3 and PT3 groups at each of the cfDNA lengths between 50 and 250bp. (B) Size profiles of cfDNA with different D-ending nucleotides across 50 and 120bp in different groups. (C) Composition of cfDNA with different D-ending nucleotides at each genomic location relative to nucleosome structure in NT2, PT2, NT3 and PT3 groups.

**Figure S4.** The concentrations of DNASE1 in the plasma of NT2, PT2, NT3 and PT3 groups.

