## Supplementary figures and images for "Detection of early-onset severe preeclampsia by cell-free DNA fragmentome"

### Supplemental Figure 1

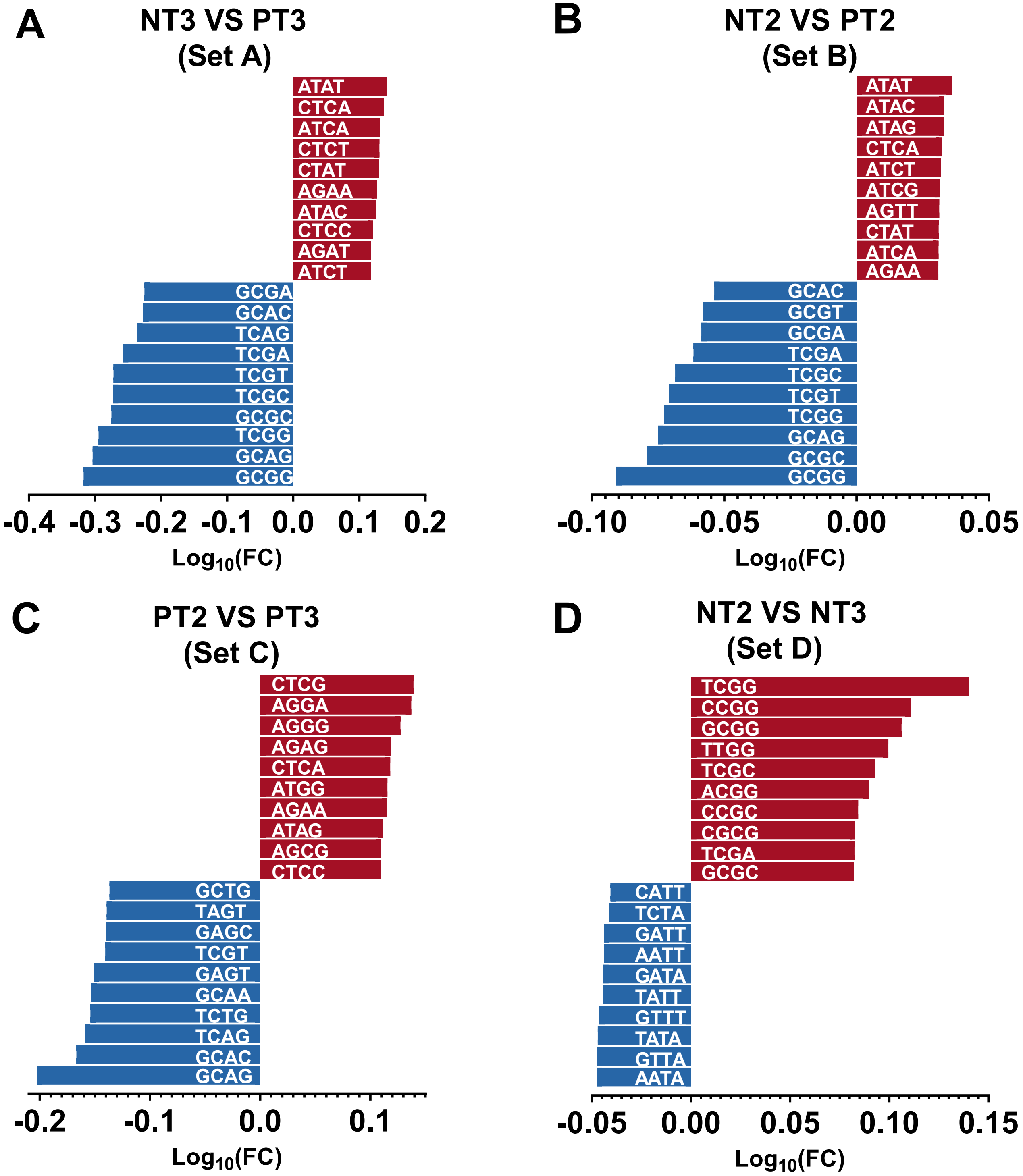

### Supplemental Figure 2

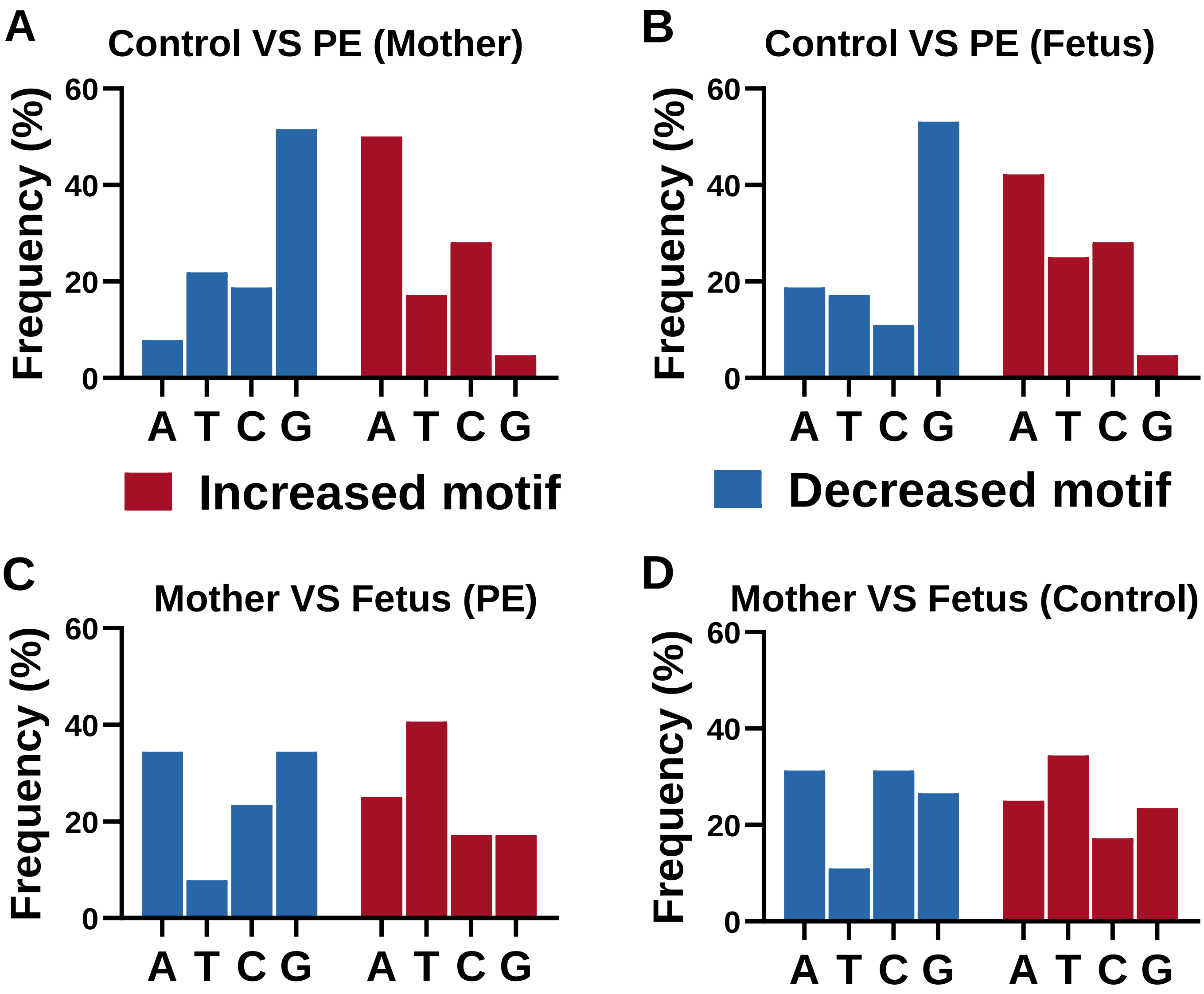

### Supplemental Figure 3

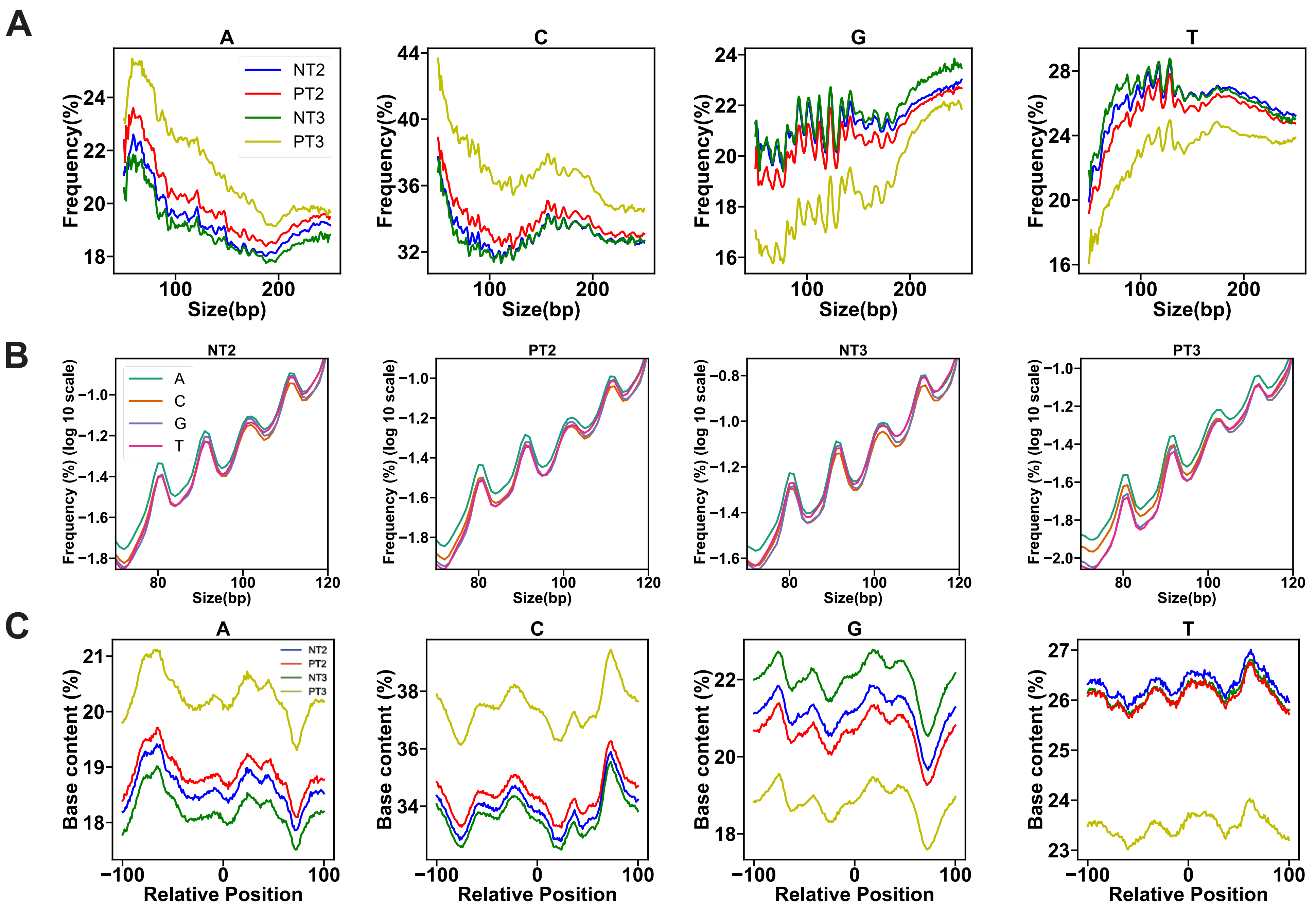

### Supplemental Figure 4

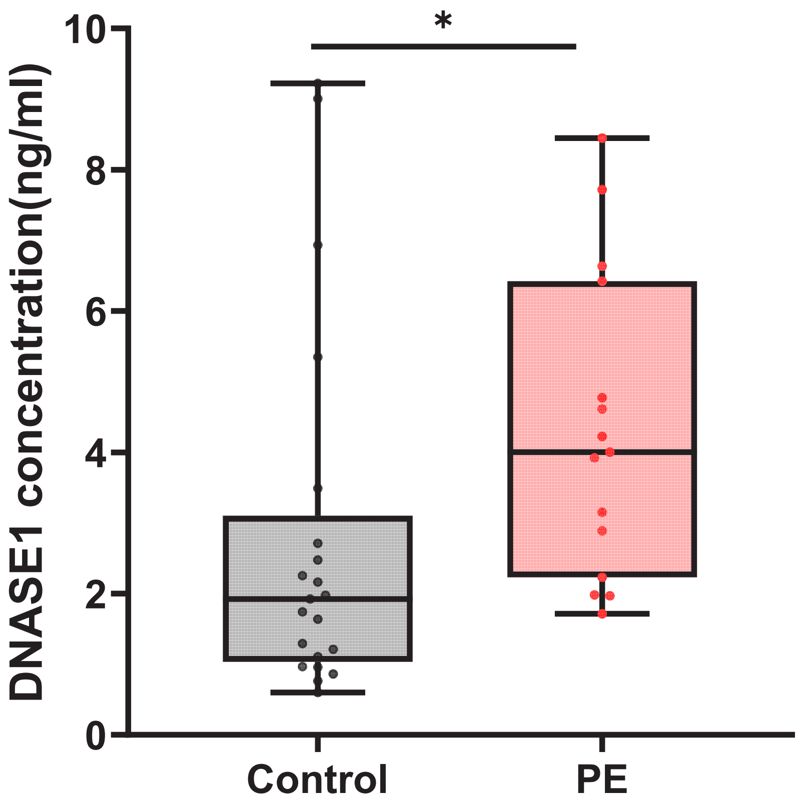
