## Supplemental Table 1 for "Detection of early-onset severe preeclampsia by cell-free DNA fragmentome"

**Supplemental Table 1. Increased and Decreased motifs idendified in discovery cohort**

| NT2 vs PT2 | | NT3 vs PT3 | | NT2 vs NT3 | | PT2 vs PT3 | |
|---|---|---|---|---|---|---|---|
| Increased Motifs n = 58 | Decreased Motifs n = 50 | Increased Motifs n = 63 | Decreased Motifs n = 64 | Increased Motifs n = 56 | Decreased Motifs n = 45 | Increased Motifs n = 63 | Decreased Motifs n = 62 |
| ATAT | GCGG | ATAT | GCGG | TCGG | AATA | CTCG | GCAG |
| ATAC | GCGC | CTCA | GCAG | CCGG | GTTA | AGGA | GCAC |
| ATAG | GCAG | ATCA | TCGG | GCGG | TATA | AGGG | TCAG |
| CTCA | TCGG | CTCT | GCGC | TTGG | GTTT | AGAG | TCTG |
| ATCT | TCGT | CTAT | TCGC | TCGC | TATT | CTCA | GCAA |
| ATCG | TCGC | AGAA | TCGT | ACGG | GATA | ATGG | GAGT |
| AGTT | TCGA | ATAC | TCGA | CCGC | AATT | AGAA | TCGT |
| CTAT | GCGA | CTCC | TCAG | CGCG | GATT | ATAG | GAGC |
| ATCA | GCGT | AGAT | GCAC | TCGA | TCTA | AGCG | TAGT |
| AGAA | GCAC | ATCT | GCGA | GCGC | CATT | CTCC | GCTG |
| AGAT | TCAG | ATAA | GAGT | CTGG | TATC | CTGG | TCAA |
| CTCC | GCCG | AGGA | GAGC | ACGC | ATTA | ATCG | GCGG |
| CTCT | TCCG | ATCC | GCCG | TTCG | CATA | AGGC | TCTA |
| AGGA | GAGT | ATAG | GCGT | CCGA | ATTT | AGAC | TCGA |
| ATCC | GCAA | ATCG | TCCG | GCGA | ACTA | ATCA | GCGC |
| ATTC | TTGC | AGAG | GCTG | GGCG | CCTA | ATGC | TCGC |
| CTAC | GCTG | ATGT | GCAA | AGCG | ACAT | ATAC | GCAT |
| ATAA | TCTG | CTTC | TAGT | AGGC | TCTT | CTCT | TCCG |
| AGAG | GAGC | AGAC | TCTG | GCCG | AATC | ATAT | TCAC |
| CTCG | TAGT | CTAA | TTGC | CGGG | CTTT | AGGT | TACG |
| AGAC | TCAA | CTAC | TTGG | CTCG | AATG | ATGT | GCCC |
| AGTC | TTGA | CTCG | TCAA | AGGG | TATG | AGAT | TCAT |
| CTAA | GCCC | AGTT | TTGA | CAGG | GTAT | CTAG | GCGA |
| ATGT | TTTG | ATTC | GCCC | ATGG | CTTA | ATAA | TATG |
| CTAG | TACG | AGTA | TTAG | ACGA | GTTC | CTGT | TTTT |
| AGTA | GAGG | AGTC | GAGG | CGGC | AAAT | CTAC | TTTG |
| ATGC | GAGA | AGGG | TACG | CCGT | TGTT | CTAT | GCCG |
| CTTC | ACGG | CTTA | TTTG | TGCG | CAAT | ATCC | GCGT |
| AGCA | GCCT | CTTT | TTGT | GTGG | TTTA | CTAA | TCCT |
| ATTG | TCCT | ATGC | GAGA | TGGG | CCTT | CTGC | TTGA |
| ATGG | TGCG | AGCA | GGGC | AGGA | AACT | AGCA | ACAG |
| AGGG | GCAT | ATGA | TCAC | CTGC | TAAT | ATGA | GAGA |
| AGCG | TCAC | CATT | TTCG | AGGT | GAAT | AGCC | TTGC |
| CTGT | TCAT | ATTG | GAAG | GGGG | TGTA | AGTC | GCTA |
| AGCT | TCTA | ATTT | TGCG | GAGG | AACA | CTTG | TCTC |
| AGTG | TTTT | CTAG | GCAT | GTCG | CATC | AGCT | GAAG |
| ATGA | GAAG | ATTA | GCCT | CAGC | CCAT | ATCT | GATG |
| AGCC | GCTC | CTGT | GCTC | GGGC | ACAA | CTGA | TTTA |
| AGGT | TCTC | ATGG | GACG | TGGC | TACT | CCGA | TCTT |
| CTTA | ACAG | CGTT | GGGG | GACG | AAAC | CTTC | GCCT |
| ATTA | TCCA | CACA | ACAG | CGGA | TACA | CCGG | GCTC |

| | | | | | | | |
|---|---|---|---|---|---|---|---|
| CTTG | TCCC | CTTG | GGGT | ACGT | TACC | AGTG | GAGG |
| CTGA | TCTT | TGAT | GGCG | TAGG | TAAC | CACG | TTGT |
| CACA | GACG | CATC | TCCT | AGAG | CCAA | CGGG | TCCA |
| AGGC | TATG | CACT | ACGG | CACG | GTAC | AGTT | GCTT |
| CTTT | GCCA | CAAT | TCAT | CGAG | | AGTA | GAAA |
| CATC | GCTA | AGTG | TTTT | CGGT | | TGGA | GAAC |
| CATT | GAAA | AGCT | TCTA | TGGA | | CCGC | ACAC |
| CAAT | TAAA | AGCC | TTAA | GGAG | | CACA | TTTC |
| GTAT | TGTG | CGAT | TAGC | ATGC | | CCGT | TCCC |
| CGTT | | AGGT | TAAG | TGGT | | CGCC | GGGC |
| CACT | | AGCG | GCCA | CCCG | | CGTC | TATA |
| TGAT | | CTGA | TCTC | CGCC | | ATTG | AAGT |
| CAAC | | AGGC | TCCC | CTAG | | CGGA | TAAA |
| CGAT | | CCTT | GGTG | CAGA | | TGAT | GATA |
| CACC | | CACC | GCTA | TGAG | | CGCG | GGGT |
| CACG | | CATA | GGAG | | | CACC | ACAA |
| CAAA | | TGTT | TCCA | | | ATTC | TAAG |
| | | CCCA | TATG | | | CACT | TGTG |
| | | CCTC | AAGT | | | CGAT | GCCA |
| | | GTAT | GATG | | | CGTT | ACAT |
| | | CAAC | ACGC | | | CCCG | GACC |
| | | CGTC | GAAA | | | AAGG | |
| | | | GTGG | | | | |

Motifs in descending order of |log(FC)|.
