## Supplemental Table 2 for "Detection of early-onset severe preeclampsia by cell-free DNA fragmentome"

**Supplemental Table 2. Validation cohort**

| | Control<br>n=48 | PE<br>n=26 | P value |
|---|---|---|---|
| Maternal age, years | 31(23-43) | 34(22-41) | 0.113 |
| BMI, $kg/m^2$ | 23.9±2.9 | 25.4±3.8 | 0.064 |
| Previous PE,%(n) | 0 | 12(3) | 0.016 |
| Times of pregnancies | 2.3±1.2 | 3.0±1.7 | 0.099 |
| Times of delivery | 0.4±0.5 | 0.7±0.7 | 0.042 |
| GA at sampling, weeks | 17.8±2.8 | 17.5±2.9 | 0.419 |
| GA at PE was diagnosed weeks | - | 31.0±2.6 | - |

PE, pre-eclampsia; BMI, body mass index; GA, gestational age.
Values were given as mean±SD and median(range).
P value: Unpaired T test (only BMI), Mann Whitney test and chi-square test.
There were cases where the number of a group may be less than the total number of the group because there was no data for individual samples.
